# Front-Line Decision-Making: A Thematic Analysis of Interviews with Hospital Staff on Referrals, Admissions, and Care for People with Multiple Long-Term Conditions

**DOI:** 10.64898/2026.02.04.26345456

**Authors:** Sara Pretorius, Sue Bellass, Rachel Cooper, Felicity Evison, Suzy Gallier, Nicola Howe, Elizabeth Sapey, Amelia Sheppard, Jana Suklan, Avan A Sayer, Miles D Witham

**Author notes:** Correspondence to: Professor Miles Witham, 3rd Floor Biomedical Research Building, Campus for Ageing and Vitality, Newcastle upon Tyne NE4 5PL.

## Abstract

**Background:** Multiple long-term conditions (MLTC) are increasingly common and place significant strain on healthcare systems designed around single-organ conditions, often resulting in fragmented and reactive care for people living with MLTC. There is limited understanding of how health care professionals (HCPs) make decisions for and with individuals with MLTC at the point of hospital presentation. This study examined how HCPs in emergency and acute settings make decisions around pathways and places of care for people with MLTC, exploring the factors that shape clinical judgement, the challenges HCPs navigate in practice and structures that influence clinical decision-making.

**Methods:** We conducted semi-structured, individual interviews with 40 NHS professionals working in emergency departments (EDs) and acute assessment units across multiple regions, roles, and specialties. Participants included consultant physicians, resident doctors, senior nursing staff and allied health professionals. Interviews focused on how decisions were made around referrals, admissions, and care planning for people with MLTC. Data were analysed thematically using an inductive approach.

**Results:** Four themes were identified: *A journey of uncertainty*, *Within and beyond limitations*, *Structures of care* and *Implementing relational care*. Clinical decision-making is shaped by clinical uncertainty, limited resources, care approaches, and interpersonal relationships and communication. Fragmented services and single-disease pathways complicate care, but participants highlighted the value of continuity, communication, and relational approaches. Challenges include resource limitations, rigid pathways and limited community support. Key enablers of clinical decision-making include integrated care, ownership, and early conversations about priorities.

**Conclusions:** Clinical decision-making by HCPs in hospitals for patients with MLTC is complex and shaped by systemic misalignment, where clinical realities clash with health system structures. Improving clinical decision-making around referrals, admissions and care planning for people with MLTC will require adapting systems and training to reflect the realities of MLTC. Potentially beneficial adaptations include strengthening relational and multidisciplinary approaches and expanding intermediate care to reduce avoidable admissions.

## Background

Multiple long-term conditions (MLTC), also referred to as multimorbidity, describes the co-occurrence of two or more chronic health conditions (1–3). MLTC are increasingly common and pose a growing challenge, not only for the individuals affected, but for healthcare systems and patient care globally (4–7). MLTC are linked to increased hospital admissions (8, 9), longer hospital stays (10), and higher risks of treatment burden and polypharmacy (10–13), exacerbating pressure on already stretched hospital systems (14). Yet hospital care remains largely structured around single-organ conditions, a model misaligned with the complexity of MLTC (15) and which may contribute to poorer outcomes (16, 17). In this context, healthcare professionals (HCPs) must make high-stakes decisions related to the care of people with MLTC within specialist environments that are poorly equipped for such complexity, often balancing competing clinical priorities where misjudgements can lead to deterioration, avoidable harm, or unplanned readmissions.

Previous research across secondary, primary, and community care settings shows that clinical decision-making around MLTC is marked by uncertainty due to physiological vulnerability, condition interactions, and limited relevant guidelines (18–22). It has been reported that HCPs often struggle to reconcile disease-specific recommendations with holistic patient needs, and existing pathways offer limited practical support (23–25). Common challenges identified by previous research include fragmented coordination, poor continuity, and limited multidisciplinary input (23, 26–28). Clinical decision-making is further complicated by the need to involve multiple professionals, within the current specialty-driven model of care, which may contribute to delayed discharge, problematic transitions and missed opportunities for integrated care (23, 25).

While existing literature has mapped the broader contextual and structural challenges of MLTC care, less attention has been paid to how decisions are made in real time at the point of hospital presentation. This qualitative study aimed to address that gap by examining how frontline hospital staff navigate clinical decision-making around referrals, admissions and care planning concerning people living with MLTC including what informs and constrains their choices and how they adapt within systems configured to provide care for single conditions. By capturing these clinical decision-making processes, we aimed for the study to provide insights that can inform understanding of current pathways and opportunities to better support person-centred, integrated care for people with MLTC, aspects that people living with MLTC consistently report as missing (23).

## Methods

### Qualitative approach and research paradigm

This study, informed by a critical realist perspective (29), used semi-structured interviews and thematic analysis (30) to explore how HCPs make decisions about referrals, admissions, and care for people with MLTC.

### Selection and recruitment

We aimed to recruit up to 40 National Health Service (NHS) HCPs across England, Scotland, Wales and Ireland, across roles and specialties, including physicians, nurses, allied health professionals, and managers working in Emergency Departments, Assessment Suites, and other acute care areas. A sample size of 40 was deemed sufficient to capture varied representation across professional roles, achieve thematic saturation, and remain feasible within the available time. Recruitment was led by the Principal Investigator, supported by the ADMISSION team, HealthTech Research Centre in Diagnostic and Technology Evaluation (HRC DTE) contacts, and the NIHR Research Delivery Network (https://www.nihr.ac.uk/support-and-services/support-for-delivering-research/research-delivery-network). Snowball sampling, outreach via the CHAIN network (https://www.chain-network.org.uk/) and study leaflets were also used. Eligible participants were English-speaking HCPs involved in secondary care decision-making for people with MLTC within the NHS in England, Scotland, Ireland and Wales.

### Data collection

One-to-one interviews were conducted online via Microsoft Teams between January and September 2024, lasting 40–80 minutes each. With consent, interviews were recorded. Participants were not reimbursed for their time.

### Interview guide

Interviews followed a semi-structured guide (Supplementary File 1), covering participants’ experiences of decision-making involving patients with MLTC, including factors influencing clinical decision-making, challenges, effective practices, and potential system improvements while allowing flexibility to explore issues raised by participants. Two pilot interviews were conducted which resulted in no change to the interview schedule. Data from these two interviews are included in the analysis.

### Patient and Public Involvement and Engagement (PPIE)

The study was co-developed with input from the HRC DTE Insight Panel, comprising patients, carers, and members of the public, and the ADMISSION Patient Advisory Group (PAG), which comprised ∼14 people living with MLTC and/or providing informal care for someone living with MLTC. The ADMISSION PAG contributed to the study’s conception and design, while the HRC DTE Insight Panel reviewed draft protocols and advised on the clarity and accessibility of participant information materials.

### Data Processing and Analysis

Recordings were transcribed using Otter.ai (v3.53.0-240624) and pseudonymised. Transcripts were reviewed for accuracy and imported into NVivo (v14) for coding. Two researchers (SP and NH) independently coded 3 initial transcripts, resolved discrepancies, and developed a shared code book which was used for subsequent data analysis and refined iteratively. Analysis followed an inductive approach, revealing meaningful narratives within the data. As analysis progressed, we explored potential differences across discipline, specialty, and career stage where these patterns surfaced inductively.

### Data Reporting

Each quotation in the results section is followed by a compact descriptor summarising the participant’s role and clinical area. To protect anonymity, consultants, consultant physicians and senior consultants are grouped under the category “consultant”. All other participants who are medically trained as doctors (e.g., specialty doctors, clinical fellows and trainees) are described using the single category “doctor”. Matrons, advanced nurse practitioners and consultant nurses are described using the category ‘senior_nurse’.

## Results

### Description of sample

A total of 40 ‘front-end’ hospital staff - those working at the initial point of hospital contact - participated in the study. Participants represented a range of clinical departments (Table 1), roles (Table 2) and locations across England and the devolved nations (table 3). Table 4 presents the role, and departmental affiliation for each participant. Participants had between 0.5 and 20 years of experience in their current role (median = 5.5 years), and 4 to 32 years in clinical practice overall (median = 18 years).

**Table 1:**
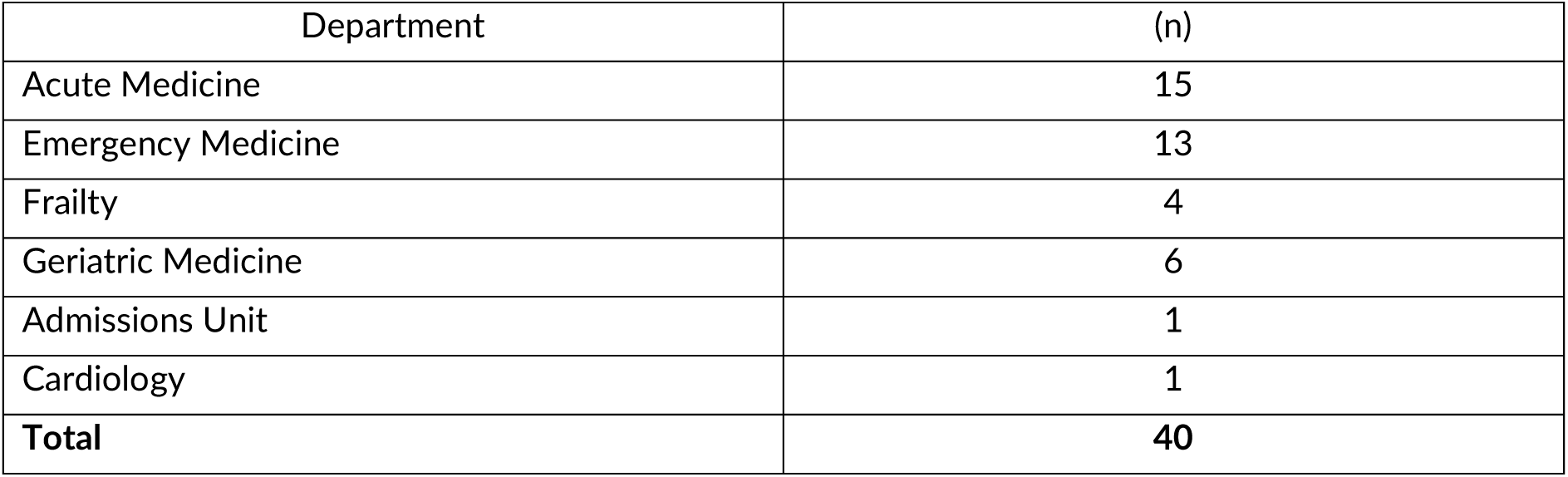
Clinical departments of study participants.

| Department | (n) |
| --- | --- |
| Acute Medicine | 15 |
| Emergency Medicine | 13 |
| Frailty | 4 |
| Geriatric Medicine | 6 |
| Admissions Unit | 1 |
| Cardiology | 1 |
| **Total** | **40** |

**Table 2:** Professional roles of study participants.

| Role | (n) |
| --- | --- |
| Advanced Clinical Practitioner | 7 |
| Clinical Academic/doctor | 4 |
| Consultant | 13 |
| Senior Nursing Staff | 3 |
| Speciality Doctor | 3 |
| Registrar | 3 |
| Manager | 2 |
| Consultant Practitioner | 1 |
| Pharmacist | 2 |
| Allied Health Professionals | 2 |
| **Total** | **40** |

**Table 3:** Geographic location of study participants.

| Location | (n) |
| --- | --- |
| Bedford | 1 |
| Birmingham | 3 |
| Bristol | 3 |
| Cambridge | 1 |
| Coventry | 1 |
| Edinburgh | 1 |
| Harlow | 4 |
| Hereford | 1 |
| Isle of Wight | 1 |
| Liverpool | 1 |
| London | 9 |
| Newcastle | 3 |
| Northern Ireland | 1 |
| Norwich | 5 |
| Nottingham | 1 |
| Plymouth | 1 |
| Salford | 1 |
| South Tees | 1 |
| Yeovil | 1 |
| **Total** | **40** |

**Table 4:**

| Participant ID | Role | Department |
| --- | --- | --- |
| ADM_Pilot_1 | Consultant | Acute Medicine |
| ADM_Pilot_2 | Consultant | Geriatric Medicine |
| ADM_1 | Consultant | Acute Medicine |
| ADM_2 | Consultant | Acute Medicine |
| ADM_3 | Consultant | Acute Medicine |
| ADM_4 | Consultant | Acute Medicine |
| ADM_5 | Consultant | Acute Medicine |
| ADM_6 | Consultant | Geriatric Medicine |
| ADM_7 | Advanced clinical practitioner | Frailty |
| ADM_8 | Doctor | Acute Medicine |
| ADM_9 | Consultant | Geriatric Medicine |
| ADM_10 | Consultant | Acute Medicine |
| ADM_11 | Doctor | Emergency Medicine |
| ADM_12 | Consultant | Acute Medicine |
| ADM_13 | Consultant | Acute Medicine |
| ADM_14 | Advanced clinical practitioner | Emergency Medicine |
| ADM_15 | Consultant | Geriatric and Acute Medicine |
| ADM_16 | Senior nursing professional | Acute Medicine |
| ADM_17 | Manager | Emergency and Acute Medicine |
| ADM_18 | Senior nursing professional | Geriatric Medicine |
| ADM_19 | Advanced clinical practitioner | Emergency Medicine |
| ADM_20 | Advanced clinical practitioner | Frailty |
| ADM_21 | Pharmacy professional | Frailty |
| ADM_22 | Doctor | Emergency Medicine |
| ADM_23 | Senior nursing professional | Emergency Medicine |
| ADM_24 | Allied health professional | Frailty |
| ADM_25 | Advanced clinical practitioner | Geriatric Medicine |
| ADM_26 | Advanced clinical practitioner | Acute Medicine |
| ADM_27 | Advanced clinical practitioner | Emergency Medicine |
| ADM_28 | Allied health professional | Emergency Medicine |
| ADM_29 | Doctor | Frailty |
| ADM_30 | Doctor | Emergency Medicine |
| ADM_31 | Consultant | Emergency Medicine |
| ADM_32 | Doctor | Emergency Medicine |
| ADM_33 | Pharmacy professional | Medicine Admissions |
| ADM_34 | Consultant | Acute Medicine |
| ADM_35 | Advanced clinical practitioner | Emergency Medicine |
| ADM_36 | Manager | Acute Medicine |
| ADM_37 | Doctor | Emergency Medicine |
| ADM_38 | Doctor | Emergency Medicine |

For anonymisation purposes, consultants, consultant physicians and senior consultants are grouped under the category "consultant". All other participants who are medically trained as doctors (including specialty doctors, clinical fellows and trainees) are described using the single category "doctor". Matrons, advanced nurse practitioners and consultant nurses are described using the category "senior nursing professional"

### Themes

Four main themes and twelve sub-themes were identified (Appendix 1). The themes capture key aspects of clinical decision-making concerning people with MLTC, including: factors influencing decisions in practice; challenges staff face when making decisions around referrals, admissions, and ongoing care; and elements of the health system that support effective clinical decision-making.

Theme 1:A journey of uncertainty

Participants highlighted the shared uncertainty experienced by both HCPs and patients at the point of their initial presentation at secondary care, particularly regarding the patient’s clinical condition, underlying health issues, suitable care pathway, and potential outcomes. This sense of uncertainty reflects the complex, evolving nature of MLTC care, where outcomes are often unpredictable due to the interplay of multiple conditions.

“We have uncertainty, we don’t know how things going to go when you have lots of diseases that are interacting with each other.” – ADM_13 (consultant_ acute_medicine)

Participant narratives indicated that decisions and actions along the care pathway are aimed at managing and reducing their own clinical uncertainty.

### 1.1 Uncertainty around diagnosis (charting territory)

For participants, the first step in reducing uncertainty is to better understand the patient in front of them. This begins with reviewing the patient’s clinical history and symptoms and conducting or reviewing investigations, to build a clearer picture of their health status. For many participants, especially those based in emergency and acute settings, the initial focus is on identifying whether the patient is presenting with an acute condition. This assessment helps determine the immediate course of action and guides the early stages of care often requiring clinicians to set aside wider considerations of MLTC in that moment.

“As an acute medic, my key skill, I find, is to unpick what’s wrong with them today, not understand what’s happened to them in their entirety” – ADM_1 (consultant_acute_medicine)

However, some participants described situations where a patient’s MLTC along with factors such as frailty status, shape referral and care decisions. At times, these factors would take precedence over referrals based solely on the acute condition. The presence of MLTC introduces additional uncertainty and often complicates a straightforward referral to a single specialist service, sometimes resulting in multiple specialist referrals to address the range of needs.

“So we would refer them to cardiology, but then, if there’s, you know, if they’re also diabetic…then we would also get a diabetic assessment…Sometimes we end up doing multiple specialty referrals” – ADM_5 (consultant_acute_medicine)

Alternatively, MLTC may preclude referral to specialist care that would otherwise be considered appropriate or beneficial to the patient.

“If that same patient wouldn’t be a candidate for a pacemaker, because they had advanced malignancy…then I wouldn’t make that referral to cardiology” – ADM_6 (consultant_geriatric_medicine)

Overall, participants highlighted the tension between the emergency focus on acute concerns and the broader presence of MLTC. In practice, HCPs described weighing immediate action against the need for longer-term planning, sometimes referring patients on to general or geriatric medicine when issues extended beyond the remit of ED or Acute Medicine. Where no acute problem was evident, decisions centred on whether conditions were controlled and home was a safe option, though the higher risk profile of patients with MLTC often tipped the balance towards admission.

### 1.2 Choosing the right companions

Participants indicated that, once a patient’s clinical status is clearer, they decide which colleagues to involve and how, whether through admission, referral, advice, or shared clinical decision-making. Participants emphasised the importance of choosing partners with both relevant expertise and a shared approach to care also noting that decisions are shaped as much by practical service engagement as by clinical fit.

“Often decisions are countered by which specialties you know will be the most helpful and which feel they have the greatest bandwidth to take on a more complicated patient. So, you know, [it’s about] who wants to go on this journey with you.” – ADM_Pilot_1 (consultant_acute_medicine)

Specialist referrals were often sought when patients required skills beyond the clinician’s own remit or where management in the community was not feasible; in cases of multiple long-term conditions, specialist input was often seen as almost inevitable. Referrals could also serve to reassure patients. While some clinicians, depending on role and confidence, were content to make decisions independently, junior colleagues highlighted the importance of involving seniors or peers when uncertainty remained, underscoring the collaborative nature of care decisions.

### 1.3 Managing risk

Participants described how perceptions of risk significantly shape their clinical decision-making, often amplifying uncertainty. They reflected on how diagnostic certainty is often pursued, not only due to clinical need but due to concerns about medical legal responsibility and accountability. Participants also described balancing the risks of discharging a patient, including the possibility of missed diagnoses, inadequate support at home, or deterioration outside of hospital, against the risks of admission, such as hospital-acquired complications, functional decline, or deconditioning. Older, frailer patients with MLTC were described as inherently more complex and at higher risk which, in turn, can influence a clinician’s willingness to assume responsibility for their care.

“People don’t say, I’m not taking them because they’re 80…but that is implicit in a lot of decisions that are made. I think the multiple long-term conditions…mean that patients are complex, and they higher risk.” – ADM_6 (consultant_geriatric_medicine)

Polypharmacy adds risk to clinical decision making for patients with MLTC, as HCPs must disentangle whether symptoms stem from underlying conditions or the effects of treatment. As one participant explained, “You need to make sure that treatment options don’t clash. Or if they have side effects…which drug or which condition or which treatment is actually causing it” (ADM_12, consultant_acute_medicine). This complexity can delay diagnostic certainty, heighten caution in treatment choices, and increase the likelihood of seeking specialist input or opting for admission rather than discharge.

Theme 2:Within and beyond limitations

The most prominent overarching theme, based on the volume of participant references, was “within and beyond limitations” which captures how constraints, both internal and external, shape the decisions made by HCPs relating to care for people with MLTC.

### 2.1 Working within a system

#### 2.1.1 Resources

Barriers to effective clinical decision-making for patients with MLTC were often tied to limited hospital capacity, staffing shortages, and gaps in essential services. These pressures are most acute in emergency departments, where the complex needs of MLTC patients clash with the fast-paced, high-turnover environment.

“When you got multiple [long term conditions], you don’t know where to start, and the [emergency] department is busy…I don’t have the bandwidth…to deal with all your 25 complaints…with an exploding department.” – ADM_19 (adv_practitioner_emergency_medicine)

HCPs described decision-making around patients with MLTC as resource-intensive and time-consuming, spanning multiple specialties and placing sustained pressure on staffing, time, and care coordination. As one participant noted, treating such patients “…involves pain teams…psychological support…multidisciplinary teams…Usually, the medics don’t like admitting these patients, because it’s very, very difficult to find a proper resolution for them. And they all become basically complex discharges.” – ADM_11 (doctor_emergency_medicine).

Participants described a sense of ‘blockage’ in the health system that constrains referral options, especially for patients with MLTC, whose complex needs often require more investigations and services, and longer hospital stays. The breadth of issues to be considered can slow decisions and compound system pressures. These blockages stem not only from in-hospital service limitations, but also from insufficient social and community care provision. This includes limited access to preventative care, a shortage of hospices and care home or community services struggling to meet the needs of complex patients.

“There’s a bottleneck at discharge…not for the simple person who had an appendix out now is going home, who’s 30 years old. It’s the people who have got uncontrolled diabetes that now need to be on insulin, but they don’t have the dexterity to do it themselves - they need a district nurse.” – ADM_38 (doctor_emergency_medicine)

Participants highlighted that resource disparities create significant inequities in clinical decision-making. Access to timely, quality care was seen to depend on local service availability and social circumstances, with deprivation and complexity compounding the challenge. A common observation was that patients living in more socioeconomically deprived areas often presented later and with more advanced illness, limiting treatment options.

#### 2.1.2 Pathways

Participants reported that decisions about where to refer patients with MLTC are often shaped by the availability of appropriate care pathways. In some cases, the chosen pathway was driven less by clinical appropriateness and more by which route enabled access to necessary investigations or management.

“There is no other pathway for getting a chest X-ray except coming through the emergency department at the moment for somebody living with frailty.” – ADM_20 (adv_practitioner_frailty)

Some participants described Trust-or site-specific admission avoidance pathways that guide clinical decision-making and support appropriate care outside hospital settings. These often promote ambulatory care or ‘home first’ models, with admission reserved for cases needing treatments that cannot be delivered on the same-day or in the community. Virtual wards, providing hospital-level care at home, were seen as key enablers of earlier discharge and safer, community-based decisions. Same day emergency care (SDEC) provides a viable option for HCPs managing patients with MLTC who require timely assessment and intervention but not admission. For HCPs unable to maintain continuity with patients, SDEC offers reassurance that appropriate investigations and follow-up will be arranged.

“Patients coming with breathlessness or chest pain come to SDEC, have an ECG, have their bloods, have a chest X-ray They are…plugged into the chest pain clinic or plugged into the respiratory clinic…If I put them on that list for a SDEC, I can sleep well.” – ADM_3 (consultant_acute_medicine)

#### 2.1.3 Time as a constraint

Participants noted that performance targets, like the NHS four-hour rule in accident and emergency departments and the 72-hour rule in acute medicine, shape how clinicians prioritise care. However, meeting such targets is especially difficult for patients with MLTC, whose complexity often demands more time and flexibility.

“We’ve got this four-hour target in A&E…within the first hour we do the examination…, investigations, and start early referrals to the right specialty. By two hours, that should be done, and by three hours someone…[asks], ‘Do you have a plan for this patient?’. But with people who have multiple conditions, it can be quite tricky.” – ADM_23 (senior_nurse_emergency_medicine)

Activities including collecting and documenting patient information, organising multidisciplinary input, and arranging investigations, were described as time-consuming in the context of MLTC. The time required for these tasks is often at odds with the realities of clinical workloads and can limit the depth and scope of assessments and decisions.

In departments not governed by time-based performance targets such as geriatric medicine, HCPs were perceived to have greater flexibility to spend time with patients and on decision making. Consultants in acute medicine were described as more likely to make rapid decisions compared with those in geriatric medicine, reflecting differing operational expectations and cultures. Long waits for specialist inputs also affect clinical decision-making. HCPs may admit patients or conduct investigations themselves to avoid delays, making admission a faster, though not always clinically necessary, route to diagnostics or treatment.

“Access to specialty probably has some impact on what I do, because I might choose to do things myself, knowing that the timeframe for getting to a specialist is too long.” – ADM_2 (consultant_acute_medicine)

Participants noted that clinical decision-making is shaped by the time of day and week, as key health and social care services are less accessible outside standard working hours (i.e. 9 to 5, Monday to Friday). This often forces staff to delay referrals or make alternative plans, especially in 24/7 emergency departments constrained by limited out-of-hours support.

“We do have a care of the elderly team…but they are not there…for the full working day…and the same goes for the navigation or the PT, OT team…so after about five o’clock, we are a bit stuck.” – ADM_11 (doctor_emergency_medicine)

### 2.2 Clinician identity and expertise

For less experienced clinicians, clinical guidelines appeared to play a significant role in shaping decision-making when caring for people with MLTC. Junior staff often relied on them for reassurance and a sense of clinical safety when navigating complex, multimorbid presentations. As one trainee doctor reflected, guidelines provided reassurance by making them feel “…more able and correct in following the step-by-step process and less likely to…make the wrong decision.” (ADM_29) (doctor_frailty). More senior colleagues described their ability to delegate clinical decision-making to more junior team members, both as a training opportunity and a necessary way of prioritising their time. They were described, or described themselves, as more comfortable with complexity, uncertainty and taking calculated risks.

“There’s some things I’m more comfortable with…I’ll manage things like heart failure, pneumonias, chronic kidney disease, frailty, cancers…I’ll do some simple tests to understand the problem better.” – ADM_1 (consultant_acute_medicine)

However, with this autonomy also comes a greater sense of liability. Consultants are seen as the ultimate decision-makers, bearing full responsibility for outcomes.

A lack of knowledge, particularly regarding community-based services, was seen to hamper timely and effective clinical decision-making, especially around discharge planning and ongoing support. Limited awareness around which community services are available for patients with MLTC reduces participants’ ability to discharge them safely, leading to delays and uncertainty in decision-making. Participants also described how outdated clinical knowledge can limit their ability to make optimal decisions concerning people with MLTC. Evolving treatments and newer care pathways were reported as often going unnoticed by more senior HCPs or those working in pressurised environments, leading to missed opportunities or inappropriate care planning.

“They’ve come from an NHS where everybody got admitted…and they’re not always keeping up to date with the alternative pathways that are now available.” – ADM_26 (adv_practitioner_acute_medicine)

Clinical decision-making also varied by clinical department and professional remit. Acute medicine staff described a focus on identifying and managing urgent needs, while also acknowledging that many of their patients have MLTC which require stabilisation as well as managing issues such as medication interactions. Emergency department staff also described an emphasis on ruling out or addressing acute problems and deciding on admission or discharge, often using structured approaches (e.g., “ABCD and everything circulation” – ADM_31). Geriatric medicine HCPs highlighted that most of their patients have MLTC and that their work centres on managing complexity. They described comprehensive geriatric assessment as core to their practice, delivered through MDT input to create personalised care plans. Once participant described comprehensive assessment as “stopping the clock” – ADM_21 on emergency care to spend more time on diagnosis. They also stressed aligning decisions with patients’ “priorities and their wishes” - (ADM_25). Frailty team members noted that frailty is not identical to MLTC but commonly overlaps.

Theme 3: Structures of care

Throughout the interviews, participants frequently referred to different structures of care representing the philosophies or approaches that shape how care is delivered and how decisions are made.

### 3.1 Organ-based care

When reflecting on the challenges of clinical decision-making around people with MLTC, many participants described a healthcare system organised around organ-specific specialisms. This was seen to foster fragmented care, relying heavily on referrals and reinforcing the separation, or siloing, of different parts of the health system.

“We no longer believe that general medicine exists. Please tell from the sarcasm in my voice that this is rubbish. But the hospital no longer believes that general medicine exists, and everyone has to belong to a speciality.” – ADM_4 (consultant_acute_medicine)

Such a specialist-driven model was considered poorly suited to the needs of a population in which MLTC are increasingly prevalent. Participants also noted that this single-organ focus makes it harder to address how multiple conditions interact or how treatment plans for one disease may conflict with another. This challenge is compounded by the persistence of single-disease frameworks and guidelines, which offer little direction for managing care in people with multiple conditions.

“We don’t have quality drivers or standards for care in people with multiple conditions, apart from the ones that sit with each single disease, which makes you think that diabetes behaves in the same way in someone who just has diabetes [compared with] someone who has diabetes, hypertension, rheumatoid arthritis…” – ADM_13 (consultant_acute/emergency_medicine)

Fragmented care makes it difficult to coordinate the right specialists. One participant described healthcare decisions as “piecemeal,” with each specialist focusing only on their own area and no-one taking an overarching view of the patient. Participants recognized the impact of this fragmentation, noting that patients often have to attend multiple appointments in different locations with some struggling to navigate this system. However, while there was an overarching sense that specialisation can fragment care, some participants emphasised that domain expertise and the availability of specialist teams often facilitates decision making by offering high-quality, targeted advice and support.

### 3.2 Joined up approach (coalescing around the patient)

Many participants advocated a more ‘joined-up’ approach, both in how HCPs make clinical decisions for patients with MLTC and in how the healthcare system itself is structured. Some participants described this more comprehensive approach as *patient-centred care*, others referred to it as *holistic care* or aligned it with the *biopsychosocial model*. While differing semantically, these terms were often used interchangeably to express a vision in which the patient is understood in the context of their whole life, not just their diagnoses.

“We look a bit more holistically at the patient, because [they may] have a social issue as well…it’s looking at the whole package of the patient, rather than necessarily what might be the presenting complaint.” – ADM_21 (pharmacist_frailty)

In this model, clinical decision-making extends beyond the acute complaint to consider a range of clinical and contextual factors including patient goals, treatment preferences, mobility, MLTC, quality of life, social context, cognitive status and psychological and spiritual wellbeing, ensuring care aligns with individual needs and capacities. Such holistic approaches were more typical among participants in geriatric or frailty roles, where comprehensive assessment is embedded in practice. This contrasts with the acute presentation focus of emergency and acute medicine clinicians, whose priority is to address an acute clinical need.

For clinical decisions to be genuinely patient-centered, participants felt it must be guided by the patient’s own wishes, expectations, and priorities, particularly for patients with MLTC.

“The main approach would be looking at…what their priorities for their life are so what they want us to treat, what they don’t want us to look into, and tailoring our approach around their preferences.” – ADM_25 (adv_practitioner_geriatric_medicine)

Participants emphasised the importance of treating patients as active partners in decision-making.

For patients with MLTC, participants emphasised that sound clinical decision-making depends on multidisciplinary collaboration. Though grounded in single-organ expertise, this approach draws together varied clinical perspectives to enable coordinated, context-aware care across physical and mental health, medication safety, social factors, and prognosis while recognising that no single HCP holds all the insight needed for safe, personalised decisions. Several participants noted that MDT access is not consistent across settings. Services with embedded MDTs, such as frailty units or specialist clinics, were perceived to support better decisions.

### 3.3 Continuous and integrated care

Continuous care is the ongoing, consistent provision of healthcare services to a patient over time and setting, typically by the same HCP or team. A lack of continuity, especially in acute settings, was seen to limit clinical options, especially when participants were unable to follow up with patients. As one emergency medicine trainee explained: “I can’t order a scan if I’m discharging them - I won’t see them again”-ADM_30 (doctor_emergency_medicine)

Continuity of care was widely regarded as essential for delivering high-quality, patient-centred care for those with MLTC, allowing HCPs to detect even subtle changes in health status and fuction, and build trust with the patient, supporting sensitive, ongoing conversations.

Clear care plans, such as advance care plans, escalation protocols and personalised management plans, support continuous care and clinical decision-making by anticipating scenarios and guiding responses. This is especially critical for patients with MLTC, where complexity can obscure the best course of action. However, such planning is often absent, leaving clinicians without a framework when health deteriorates rapidly.

“Advance care planning is one of the biggest things, and often the lack of that comes about from people who’ve maybe not had many interactions with the health service, [who have] quite quickly…become multi-morbid” - ADM_14 (adv_practitioner_emergency_medicine)

Having plans in place from the start also ensures a shared understanding across services and settings.

Participants stressed the importance of intermediate care resources, such as step-up or step-down units in making decisions which support care continuity. These allow patients to receive the care they need without occupying an acute hospital bed.

“One of the things we’re looking at that may be helpful is increasing our intermediate care beds You don’t need to stay in that acute bed, but going to an intermediate care bed will be much more beneficial.” – ADM_10 (consultant_acute_medicine)

Continuity of care for patients with MLTC includes appropriate follow-up and support after discharge. Decisions about sending patients home hinge on available community support including healthcare, social care, and family involvement, often described as a ‘safety net’ to prevent gaps in care. However, access to community health and social care services varies widely, with many participants facing limitations. As one Acute Medicine clinician expressed “If you could have a magic wand to make one thing better…it would be the community provision of care”. Other participants reported strong local systems.

“In terms of managing patients with these multiple comorbidities, we know what services are available for them, particularly for us in the community setting..having community dietetics or the community physio.” – ADM_18 (senior_nurse_geriatric_medicine)

These discussions underscored how the presence of supportive community care fosters a sense of security and confidence in clinical decision-making. The contrasting accounts may partly reflect disciplinary perspectives: acute medicine clinicians, who frequently face gaps in community provision when planning discharge, contrasted with geriatric teams—more embedded in community pathways who often have greater awareness of available services.

Participants emphasised that continuity of care for patients with MLTC relies on integrated decision-making across services including platforms or processes that enable teams from general practice, social care, mental health, and secondary care to coordinate around patient needs. In-reach models were seen as particularly valuable, allowing specialist teams to contribute within the patient’s current setting rather than relying on referral, thereby streamlining decisions, reducing disruption and reassuring clinicians that patients would receive the necessary multidisciplinary care.

Theme 4:Implementing relational care

The theme ‘implementing relational care’, highlights the role of strong relationships in clinical practice and decision-making. These connections between HCPs, patients, families, and other professionals, help navigate competing demands and shape the quality of care.

### 4.1 Someone to take responsibility

Participants described the difficulty of securing specialist clinicians to take responsibility for the care of patients with MLTC.

“So if you’re unsure of what is driving the problems, or you think it might be a combination…of things, it is very difficult to get an ology-focused consultant to…take ownership and leadership.” – ADM_Pilot_1 (consultant_acute_medicine)

Often specialists perceive multiple conditions as the domain of ‘generalist’ physicians tending to view their role as confined to a single condition, and therefore not as the appropriate person to take a broader perspective on the patient’s overall care. When departments or specialists decline to accept responsibility for patients, it not only creates challenges for those making referrals but also prolongs care pathways and contributes to system blockages with clinicians “spending all day battling between who feels they should take responsibility for the patients.” – ADM_27 (adv_practitioner_emergency_medicine).

Participants highlighted the importance of having confident, experienced decision-makers, particularly senior clinicians who can navigate uncertainty, assess complex cases quickly, and take responsibility for key decisions around admission, discharge, and ongoing care for patients with MLTC.

“What they actually need is a named consultant who can be involved in their care and make a judgement call as to ‘Yes, this is appropriate’ or ‘this isn’t’.” – ADM_4 (consultant_acute_medicine)

Participants highlighted the importance of patients taking responsibility for their own health. For patients, this engagement fostered a tangible sense of empowerment and control, supporting improved monitoring and preventative measures. Such patient-led action was seen to help prevent deterioration to the point of requiring emergency care, thereby alleviating pressure on emergency services and reducing barriers to effective clinical decision-making. Patient-led self-care was considered a facilitator of effective clinical decision-making for MLTC. When patients are empowered to manage their health, they contribute valuable insights that shape treatment choices.

“Giving people ownership and flexibility to look after the condition makes them so different…They can tell you actually, I’ve tried this drug before, it doesn’t work.” – ADM_12 (consultant_acute_medicine)

### 4.2 Conversation and communication

Meaningful conversations with team members, colleagues in other departments, the multidisciplinary team, community partners, patients, and their families, were seen as essential for implementing relational care for people with MLTC. Participants highlighted the importance of having trusted ‘go-to’ contacts within specialist services and forming strong communication links with staff in other departments and community services in facilitating clinical decision-making. Interviewees working in smaller hospitals noted that the physical proximity of departments made it easier to have face-to-face conversations with colleagues from other specialties leading to faster collaboration and joint decision-making.

Access to reliable information was seen as crucial for relational care, enabling informed discussions and shared decisions. Yet participants expressed frustration with fragmented communication systems, citing poor documentation, lack of continuity, and incompatible IT platforms, often leaving teams without essential data or reliant on incomplete, inconsistent records.

“Information sharing is probably one of the biggest factors in that…it’s just very sporadic and very patchy.” – ADM_14 (adv_practitioner_emergency_medicine)

These limitations hindered a full understanding of patients’ histories, conditions, and contexts. Participants reported wide-ranging consequences including repeated tests, treatment delays, inappropriate admissions, and medication errors, especially affecting those with MLTC.

### 4.3 Balancing priorities and managing conflict

Clinical decision-making for people with MLTC is influenced by the HCP’s ability to balance competing demands, whether from different individuals or services, or inherent in the care itself, a challenge that is heightened in MLTC, where multiple conditions and stakeholders create layered and often conflicting priorities. Conflicts may arise, for example, when the priorities and expectations of patients and their families differ. As one participant noted, “The problem you often have is, what the patient wants isn’t necessarily what the family wants. And…you can [only] win some battles.” – ADM_1 (consultant_acute_medicine).

HCPs and patients may also disagree, particularly about whether certain interventions are appropriate given the patient’s overall health and frailty. Such disagreements may even lead clinicians to decline accepting a patient when they feel unable to meet the expectations placed upon them.

“If there’s a patient who’s 95 years old, and they insist they want to be for CPR, and they’ve got chronic health conditions, and they look so frail…I don’t [accept them as a patient].” – ADM_1 (consultant_acute_medicine).

Participants also described the tension within multidisciplinary teams which can complicate clinical decision-making, particularly when different professionals or services hold divergent views.

Beyond interpersonal dynamics, participants also spoke of having to balance the needs of their patients with the constraints of the wider healthcare system. This tension was further reflected in frustrations about the gap between the care clinicians wanted to provide and what was practically achievable, and the resulting impact this has on relationships with patients.

“Sometimes it’s been as brutal as kind of saying, okay, so this is what I can do today. This is what you want us to do. I can’t do X and Y…What would be a compromise?” – ADM_35 (adv_practitioner_emergency_medicine)

Ultimately, the tension between what constitutes ideal care and what is clinically feasible leaves many participants grappling with difficult trade-offs. The necessity of making such decisions often carries a moral and emotional weight, with HCPs describing feelings of frustration, demotivation, and even moral injury when constrained by systemic limitations.

## Discussion

This study explored how HCPs in emergency and acute settings make decisions with and for people with MLTC who attend hospital, focusing on referrals, admissions, and patient care planning. Four themes captured their perspectives: a journey of uncertainty, within and beyond limitations, structures of care and implementing relational care.

Clinical decision-making is shaped by the uncertainty of managing acute illness in the context of MLTC, requiring HCPs to balance clinical risk, judge whether to prioritise acute needs or broader health concerns, seek support from knowledgeable colleagues, and stay focused on the aims of the visit. HCPs operate within constraints such as system pressures, limited resources, patient expectations, and rigid time targets, all of which influence the speed, depth, and nature of clinical decision-making. Services structured around single conditions, fragmented discharge pathways, and insufficient community or social care capacity add further complexity. Participants from geriatric medicine and frailty settings described adopting a more holistic approach, considering the wider context of the patient, including social circumstances and life stage, whereas those in acute and emergency medicine, though mindful of MLTC, often feel compelled to prioritise the acute clinical needs.

Participants highlighted the need for more integrated approaches that promote continuity, clearer ownership, and earlier discussions about goals of care for people with MLTC. Relationships, with patients, families, and colleagues, are central to how decisions are made, supporting communication, shared understanding, and coordinated care.

### Findings in relation to previous research

HCPs described clinical decision-making around people with MLTC as highly uncertain, shaped by unclear presentations and pathways. Actions such as ordering additional investigations aimed to reduce uncertainty but rarely eliminated it, reflecting previous research highlighting the complexity, physiological vulnerability, and lack of clear guidelines in MLTC decision-making (18–22). Consistent with previous research, MLTC was reported to create complex, fragmented care challenges that strain standard pathways and health system resources, complicating clinical decision-making and limiting available options. (20, 24, 26). Continuity emerged as a key factor shaping clinical decision-making, with longitudinal relationships supporting trust, early recognition of deterioration, and informed advance care planning. This aligns with prior evidence linking continuity to improved patient outcomes and satisfaction (21). However, continuity is often disrupted in acute care, underscoring the critical role of primary and community services in enabling HCPs to make timely, coordinated, and holistic decisions (26). Integrated models, including specialist in-reach teams, virtual wards, and step-up/step-down units, were seen as important for bridging hospital–community gaps, providing HCPs with more care options and flexibility in clinical decision-making (27, 31).

Clear ownership of patient care also emerged as vital, with generalist services often assuming responsibility for complex cases. Patients and families were seen as active clinical decision-making partners, consistent with literature on the importance of empowerment and engagement in MLTC management (32, 33). HCPs described navigating competing demands, including those between acute and chronic needs, and ideal care versus organisational constraints. These ethical and practical tensions have been noted previously in MLTC care (19). In alignment with previous studies, our findings suggest that multidisciplinary collaboration, underpinned by communication and negotiation skills, is essential to managing such conflicts (24, 31). Our findings around the constraints of limited resources and systemic pressures align with previous research, which has highlighted how workforce shortages, time pressures, and fragmented funding structures restrict the delivery of coordinated, patient-centred care for people with MLTC (19, 24, 25, 27). Our findings also align with recent recommendations from UK health policy think tanks to strengthen care for people with MLTC. The King’s Fund highlights the need for holistic, coordinated care, clearer responsibilities among those involved in MLTC management, and improved digital infrastructure (34). Separately, Re:State proposes establishing a new generalist hospitalist specialty to provide clinical leadership for patients with undifferentiated, complex needs across hospital and community settings (35).

### Strengths and limitations of the current study

A key strength of this study is its direct focus on clinical decision-making, offering fresh insights into how HCPs in acute and emergency settings navigate the complexity of delivering health care for patients with MLTC. The study also draws on a large and diverse sample of 40 participants across varied roles, specialties, and locations, capturing both shared challenges and distinct, context-specific perspectives.

Some limitations should be acknowledged. Interviews reflect reported rather than observed practice, meaning some aspects of real-time clinical decision-making may be under-or over-represented. While we included a range of professional groups, this study did not capture patient or carer perspectives, which would offer valuable complementary insights into clinical decision-making. However, these perspectives have been explored within the ADMISSION research programme through a recent qualitative study examining the lived experience of inpatient hospital care among people living with multiple long-term conditions (MLTC) [in preparation]. There are clear parallels between the findings of the patient-focused study and our clinician-focused analysis. Both identify persistent tensions between care for acute and chronic conditions within a predominantly specialist model of hospital care, resulting in uncertainty in care pathways. A further disconnect emerges between the complexity of patients’ needs and the organisation of care around single-condition approaches, reflected in the continued reliance on single-condition clinical guidelines (22). These challenges are navigated within inflexible models of care and resource-constrained hospital settings, shaping experiences of hospital care for people living with MLTC and clinicians’ capacity to make holistic decisions. Across both studies, integrated care, effective inter-and intradepartmental communication, and generalist clinical knowledge emerge as important in supporting holistic and efficient clinical decision-making and care within hospital systems.

### Directions for future research

Future work should explore how to improve health system structures and processes based on the challenges and suggestions raised by HCPs in this study, particularly around system constraints, communication, and support for clinical decision-making. Ethnographic and observational methods could further enrich understanding by capturing the real-time dynamics of clinical decision-making in acute and emergency care. Comparative studies across healthcare systems with different models of integration, funding, and resourcing could also illuminate how structural factors support or constrain clinical decision-making for MLTC.

### Implications for Practice, Policy, or Theory

This study has important implications for practice, policy, and theory in the care of people with MLTC. Practically, there is a clear need to strengthen training for HCPs in MLTC management, especially through experiential and inter-specialty learning, to better equip clinicians for the complexity of MLTC in acute and emergency care.

Policy-wise, investment in integrated, flexible, and well-resourced services is critical if integrated care boards and related initiatives are to fulfil their potential. Theoretically, our results reinforce the need for collaborative, systems-based models of clinical decision-making, emphasising the role of continuity, context, and teamwork. Our findings also inform workforce planning, pointing to the value of mentorship and exposure to complex clinical decision-making for junior staff. Supporting clinician wellbeing and retention may depend on recognising the interpersonal and adaptive skills that underpin clinical decision-making and care for people with MLTC.

## Conclusion

Clinical decision-making for people with MLTC remains complex and is often constrained by systemic misalignment, where day-to-day clinical realities sit uneasily within existing service structures. Strengthening decision-making around referrals, admissions and care planning will require systems and training that better reflect the realities of MLTC. Promising directions include reinforcing relational and multidisciplinary ways of working, improving coordination across services, and expanding intermediate and community care to reduce avoidable admissions and ease pressure on hospitals.

## Supporting information

Interview schedule

## List of abbreviations

Abbreviation: Non abbreviated word/phrase
A&E: Accident and Emergency
AI: Artificial Intelligence
CHAIN: Contact, Help, Advice and Information Network
CPR: Cardiopulmonary Resuscitation
ED: Emergency Department
HCP: Healthcare professional
HRA: Health Research Authority
HRC: DTE HealthTech Research Centre in Diagnostic and Technology Evaluation
IT: Information Technology
MDT: Multidisciplinary Team
MLTC: Multiple long-term conditions
NIHR: National Institute for Health and Care Research
OT: Occupational Therapy
PPIE: Patient and Public Involvement and Engagement
PAG: Patient Advisory Group
PT: Physiotherapy
REC: Research Ethics Committee
SDEC: Same Day Emergency Care

## Declarations

### Ethics approval and consent to participate

Interested individuals received a Participant Information Sheet and consent form by email. Those who agreed to participate returned a signed form or gave verbal consent, prior to interview. The study was approved by the NHS Health Research Authority and Health and Care Research Wales (REC reference: 24/HRA/1483) and conducted in line with the Declaration of Helsinki (2008).

### Consent for publication

This manuscript does not contain any individual person’s data in any form (including identifiable details, images, or videos). Written informed consent for publication of pseudonymised quotations was obtained from participants, as appropriate.

### Availability of data and materials

Pseudonymised interview data used in this publication which are not provided in the supplementary materials are available from the corresponding author upon request or from the Newcastle University Figshare repository (https://doi.org/10.25405/data.ncl.31198234).

## Authors’ contributions

MDW, SB, SP, JS and NH were responsible for the study conceptualisation. SP, NH and AS were responsible for data analysis. SP was responsible for initial manuscript writing, NH, SB, MW and JS were responsible for initial manuscript review. All authors were responsible for the review and editing of the manuscript. MDW is the guarantor for this work. The corresponding author attests that all listed authors meet authorship criteria and that no others meeting the criteria have been omitted.

AAS, RC, ES and MDW acquisition of funding.

## Appendix 1: Themes, subthemes and code clusters

**Theme 1.**
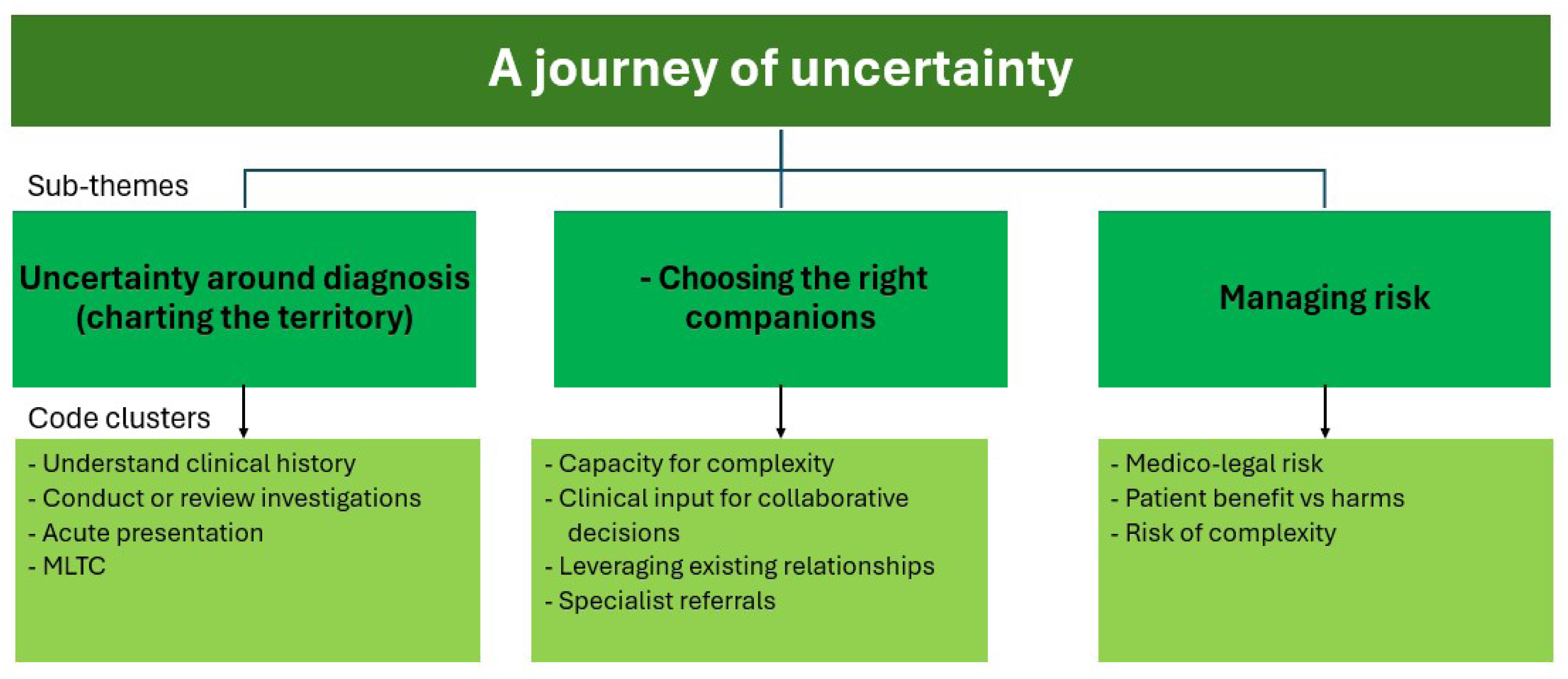

**Theme 2.**
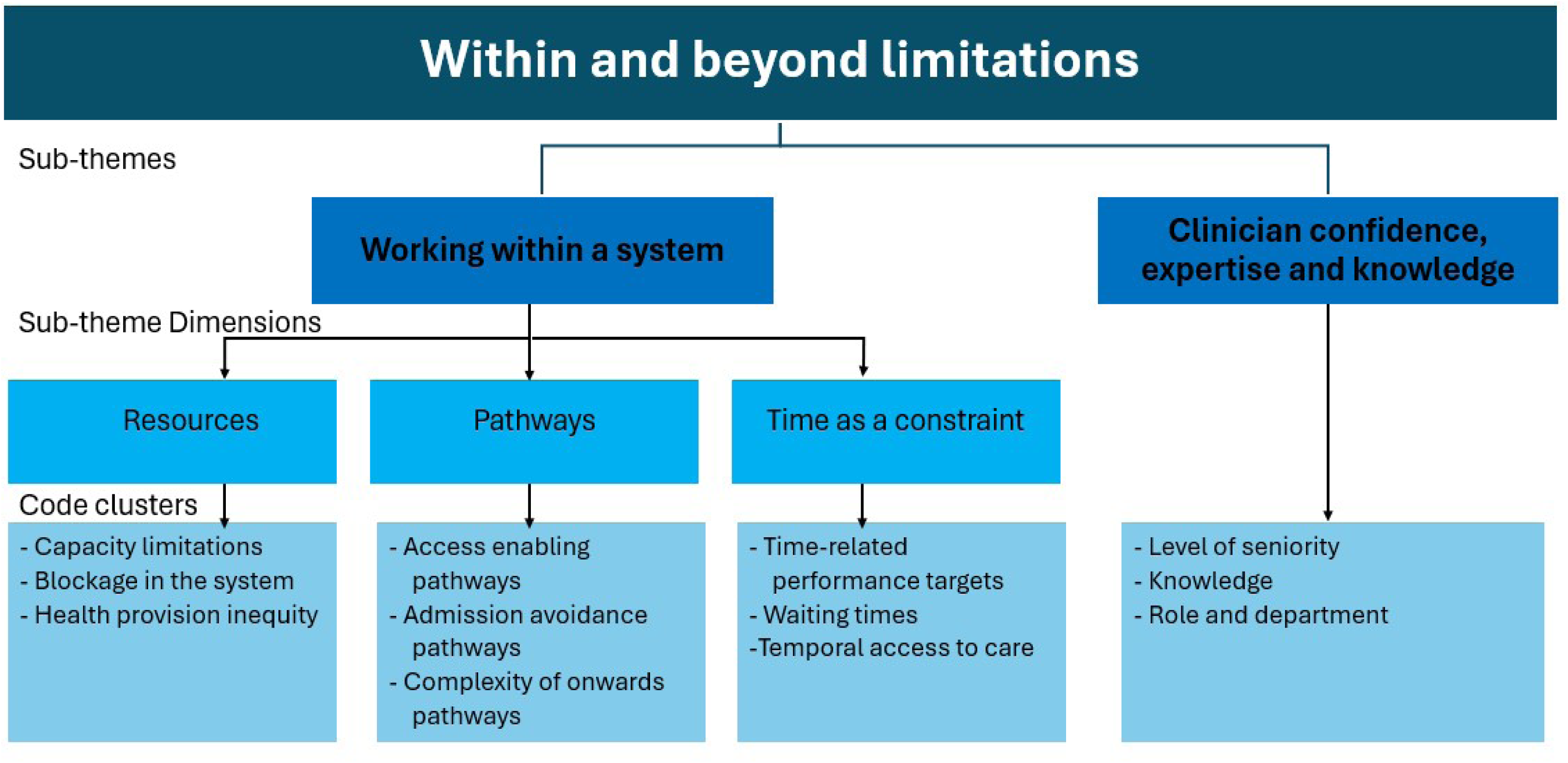

**Theme 3.**
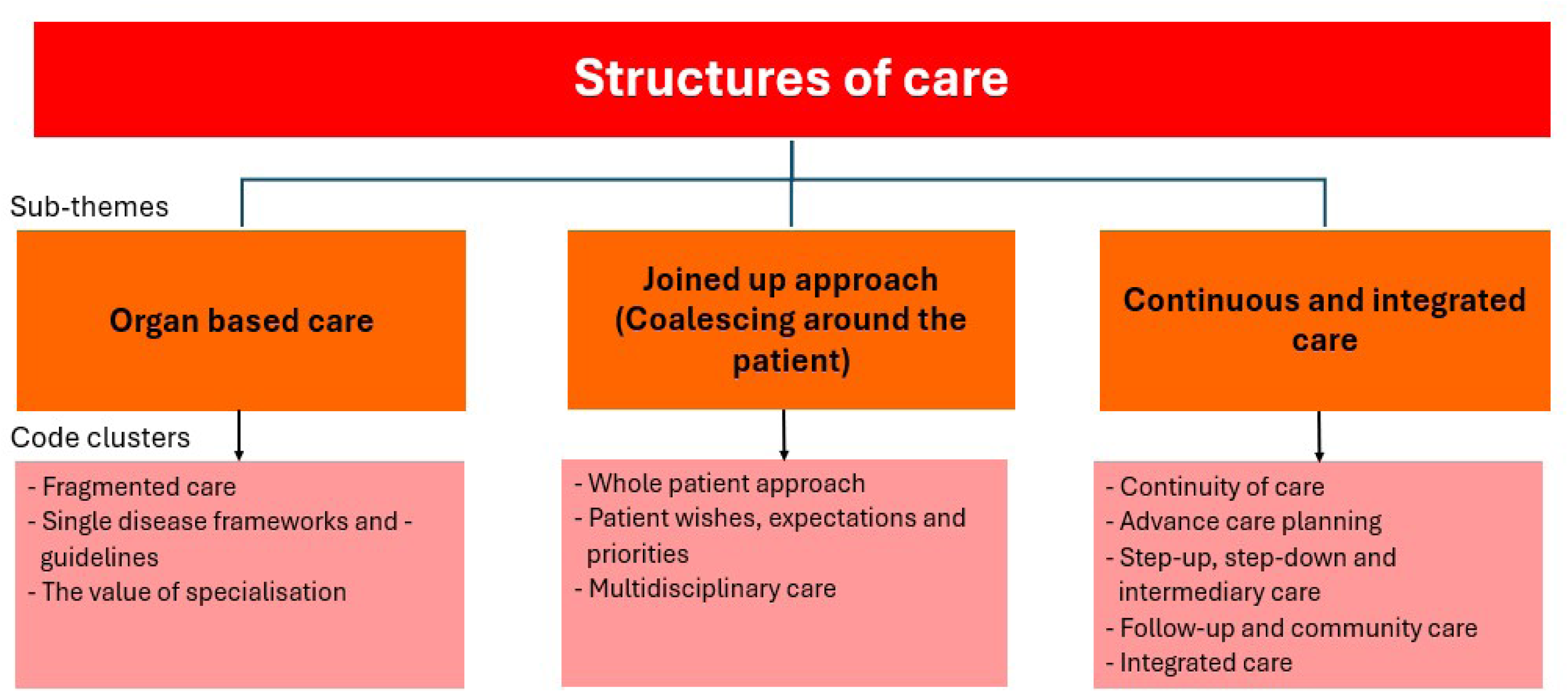

**Theme 4:**
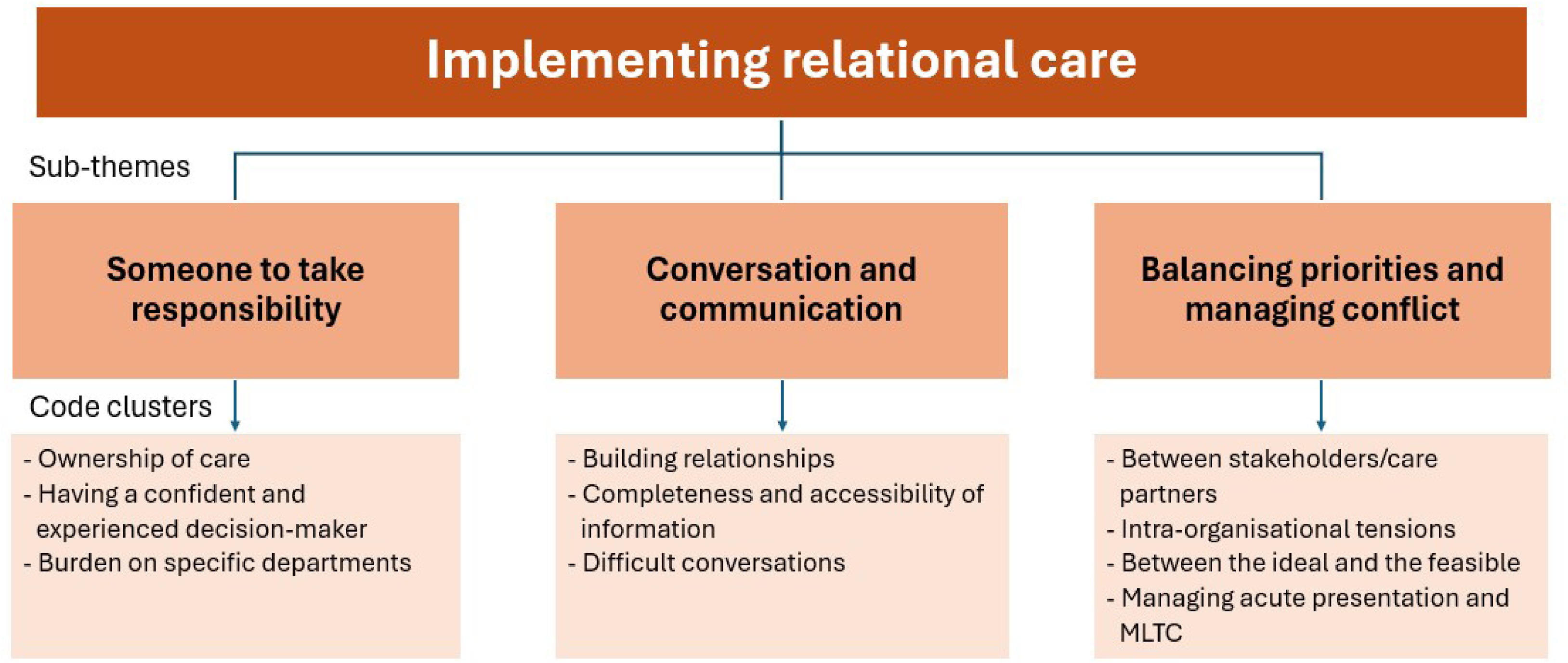

