## Supplementary material for "Front-Line Decision-Making: A Thematic Analysis of Interviews with Hospital Staff on Referrals, Admissions, and Care for People with Multiple Long-Term Conditions": Interview schedule

### Supplementary File 1: Interview Schedule

### Clinical decision-making and patient pathways for patients with multiple long-term conditions (MLTC) in secondary care

#### ADMISSION - CPA

#### Clinician Interview Schedule

#### Background

The Newcastle HRC team will perform a maximum of 40 Interviews with NHS hospital staff in different roles and specialties (nurses, consultants, ACPs, physiotherapists, pharmacists, managers) working in the:

- Emergency department (ED)
- Acute medicine department
- Other acute assessment areas

These interviews will explore the following themes:

- demographic information
- clinical decision-making around referral and management of patients with MLTC.
- Factors influencing clinical decision-making for patients with MLTC
- challenges and strengths of the current system for managing patients with MLTC

An interview schedule has been developed below, which will be piloted during the initial phase of the study.

### Interview Schedule

#### Introduction

‘Thank you for taking time for this interview.’

‘This will be a semi-structured interview, but before I start asking you questions, let me give you a brief overview about who we are.

‘My name is [INSERT] and today we are also joined by [INSERT], and [INSERT].

‘Today we would like to discuss with you the how you make decisions around admissions, referrals and the management of patients with MLTCs coming to the hospital through the Emergency Department or Acute Assessment Suite. Our aim is to understand current practice in clinical decision-making for people with MLTC in hospitals.’

Have you had time to review the information sheet?

Do you have any questions?

[TAKE THROUGH CONSENT PROCESS]

‘Do you have any other questions before we proceed with the interview?’

#### Questions

**START RECORDING**, if consented to.

#### Demographics

- Where are you based?
- What is your current role within the hospital?
    - What sort of hospital do you work in? (teaching/research, community specialist centre, or smaller practice)
- How many years of experience do you have in your current role?
- When did you qualify for clinical practice?
- Where did you qualify?
    - UK, overseas?

#### Current Practice

- Can you describe the typical approach in your speciality for making-decisions around patients with multiple long-term conditions particularly in terms of admissions, referrals and care management?
- What do you perceive as the current challenges or limitations when making decisions for patients with MLTC?
- What works well within the current system for making decisions around patients with MLTC?
- What factors influence your decision-making around patients with MLTC?
    - Organisational culture, resources/capacity limitations
- What changes or improvements are needed to make the system more efficient for making decisions around MLTC patients / clinicians - How do you think this can be achieved?

#### Closing comments

- Is there anything else you feel is important to tell us about managing patients with MLTC that we haven't touched upon in this interview?
- Do you have any feedback on the interview?
